# Incidences of poor-quality pharmaceutical products in Nepal

**DOI:** 10.1101/2021.04.15.21255541

**Authors:** Astha Neupane, Maheshwor Bastakoti, Sabita Tamang, Basant Giri

## Abstract

Pharmaceutical products are used to treat, prevent, and save lives of millions of people globally. However, pharmaceutical products known as substandard and falsified that do not meet regulatory standards and quality threaten the health of the population of today and future leading to socio-economic hardship, drug resistances and put life of patients in danger. We analyzed the recall notice from 2010 to 2020 issued by the department of drug administration (DDA), government of Nepal to understand the overview of substandard and falsified pharmaceutical products in Nepal. The number of recalled pharmaceutical products has significantly increased over the past decade in Nepal (p-value< 0.05). The most recalled drugs were antimicrobials followed by gastrointestinal medicines, vitamins and supplements, pain and palliative medicines among others. Number of recalled drugs manufactured by domestic pharmaceutical producers and imported ones were not significantly different. Majority of imported recalled drugs originated from India. Sixty-two percentage of recalled drugs were substandard, 11% were falsified and remaining 27% were not registered at the DDA. Similarly, sixty percentage of recalled drugs were modern and 35% were traditional ones. The hand sanitizers used to minimize the COVID-19 transmission contributed significantly to the list of recalled pharmaceutical products in 2020. Most of these sanitizers contained significant amount of methanol instead of ethyl alcohol or isopropyl alcohol. We also reviewed peer-reviewed research papers that reported data of substandard and falsified drugs. Only four such papers were found in literature. These papers reported issues with labeling, unregistered drugs and drugs failed in a number of laboratory testing. Since the recall data did not include number of samples tested and location of sample collected, a systematic study to understand the prevalence of substandard and falsified drugs in Nepal is recommended.

**Key questions:** *What is already known?:*

- Prevalence of substandard and falsified pharmaceutical products is a global threat to public health and socio-economy.

*What are the new findings?:*

- We analyzed drug recall data from department of drug administration in Nepal and report that the substandard and falsified pharmaceutical products are increasing significantly.
- Antimicrobial drugs were the most frequently recalled drugs. Drugs manufactured by domestic producers and imported ones were equally recalled. Allopathic drugs were recalled more than ayurvedic products.

*What do the new findings imply?:*

- This study suggested the problem of substandard and falsified pharmaceutical products is serious in Nepal. Integrated efforts from regulating agencies, manufacturers and pharmacies are recommended to minimize the circulation of such products in the country.

## Introduction

Pharmaceutical products are essential products used to treat, prevent, and save lives of millions of people globally^1^. They should be safe, effective, and of good quality. Such products should be prescribed and used rationally^2^. However, pharmaceutical products that do not meet regulatory standards and quality threaten the health of the population of today and future. Low-quality drugs could lead to drug resistances and put life of patients in danger in addition to creating economic and social burden to people^3^. There are several reasons for the circulation of such substandard products in market such as lack of access to affordable, quality, safe and effective medical products, and good governance, poor ethical practices in health care facilities and medicine outlets. Limited technical capacity in manufacturing, quality control, and distribution also contribute to the same problem^4^.

A recent meta-data analysis estimated that about 10.5% of the medicines worldwide are either substandard or falsified. Prevalence of low-quality pharmaceutical products is higher in low- and middle-income countries (13.6%) compared to high income countries. About 18.7% medicines were estimated to be low-quality in Africa and 13.7% in Asia. The most substandard or falsified drugs were the antimalarials (19.1%)^3^.

Nepal is one of the least developed countries that shares open and poorly regulated boarders with India and China. These two countries are considered as the major producers of low-quality and fake pharmaceutical products circulating in the global market^4^. The domestic market for medical products in Nepal was estimated to be 70 billion Nepal rupees in 2019 that included drugs (36 billion), raw materials and surgical equipment^5^. The department of drug administration (DDA), which is the drug regulatory body of Nepal is responsible to authorize the distribution of all pharmaceutical products. The DDA also aims to prevent the misuse or abuse of drugs and allied pharmaceutical substances, the false or misleading information relating to the efficacy and use of drugs and, to control the production, sale, distribution, export, import, storage and consumption of those drugs which are not safe, efficacious, and of standard quality^6^. Few studies in past have indicated the circulation of substandard, counterfeit, and unregistered drugs in the Nepali market^7,8,9^. The DDA Nepal recalls marketed drugs if the drugs do not fulfill any requirement as indicated in the drug act 2035 B.S.^6^. It then issues public alerts and warnings when substandard, falsified and unregistered medicine incidents are detected.

In this study, we analyzed the incidences of poor-quality drugs in Nepal. We used drug recall notice issued by the DDA and research publications. We analyzed temporal trend of low-quality drugs, types of drugs and formulations. We report the low-quality drugs have increased significantly in Nepal that over the last decade and among them antimicrobials are the most commonly found low-quality drugs.

## Methodology

We carefully analyzed drug recall notice published by DDA Nepal from 2010 to 2020. The DDA publishes such notices in its bulletins, websites and newspapers. We extracted important information provided on the recall notice such as brand name, dosage form, batch number, manufacturing date, expiry date, recall date, reason of non-compliance, and the manufacturer information. We used National List of Essential Medicines 2016 of Nepal to classify the recalled drugs into essential and non-essential drugs^10^. We used the WHO definition to identify substandard, falsified and unregistered drugs^11^. According to WHO definition, the substandard drugs are authorized medical products but fail to meet quality standards or specifications or both. Similarly, falsified drugs are medical products that misrepresent their identity, composition or source^12^. Pharmaceutical products that did not pass dissolution test, API assay, microbial test, leakage test, friability, non-compliance with pharmacopeia, physical appearance, fungal count, weight variation, particulate matter test, uniformity test, disintegration test, and pH test were put together under substandard category. Similarly drugs that contained impurities, active ingredient not meant to be there, had price sticker without approval, and did not have product specification were classified as falsified pharmaceutical products. The drugs that were recalled because they were not registered at DDA Nepal were classified under unregistered category. Unregistered drugs do not undergo evaluation and/or approval by DDA Nepal. We classified the drugs as “others” if the reason of recall was not specified.

In addition to recall notice, we also analyzed peer reviewed research articles from electronic databases such as PubMed (2010-2020), Web of Science (2010-2020), Springer link (2010-2020), and Google Scholar (2010-2020). We used the following search terms in conjunction with Boolean search term (“OR”, “AND”) to identify related articles: “counterfeit*”, “substandard*”, “falsified*”, “fake”, “spurious”, “unregulated drugs”, “unregistered”, or “frauds”; combined with “Drug”, “medicine”, or “pharmaceutical”; “Nepal*”. In Google Scholar same search term was used, but instead of “Nepal*”, we used “intitle:Nepal”. The articles were screened and evaluated manually through the title and abstract on the basis of inclusion criteria: date of publication (2010-2020), the language (English) in which the article was published, the article should contain data/information on the prevalence of falsified/spurious/counterfeit/substandard drugs and the location of experiment/research carried out. Similarly, the articles which did not meet inclusion criteria were excluded. We also did not include opinion articles, letters, notes, conference papers, book chapters, editorials or comments or articles with no abstracts or articles with counterfeit or substandard medicines related to animals.

Statistical analyses of data such as Chi-square test, Fisher exact test and simple linear regression were performed using R version1.4.1106.

## Results

We analyzed recalled drugs during 2010 – 2020. During this period 346 pharmaceutical products were recalled by DDA Nepal. The number of recalled low-quality drugs in Nepal has significantly increased in the last decade (figure 1A, linear regression, p value< 0.05, adjusted R-squared value= 0.335). We found that only one pharmaceutical product was recalled in 2010. The product was the lactate solution which is commonly used for fluid resuscitation. The lactate was recalled from Nepali market since it did not pass the sterility test. There was no recall in 2012. The year of 2018 had the highest number of pharmaceutical products recalled (123 products, see figure 1a). Forty-six products were recalled in the year 2020, majority of which were hand sanitizers. The recalled pharmaceutical products were from 96 manufacturers mostly from Nepal and India, few from Bangladesh, and China. Manufacturer of 91 recalled drugs were unknown.

**Figure 1:**
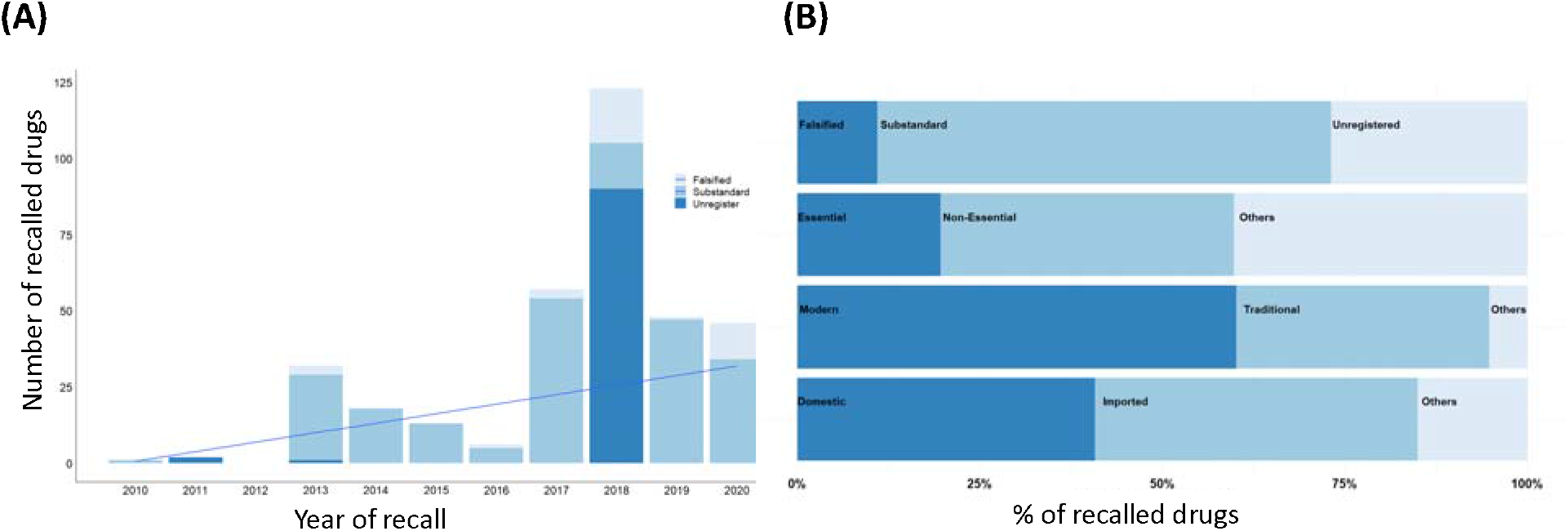
(A) Temporal trend of recalled pharmaceutical products in Nepal. (B) Contribution of different category of pharmaceutical products in the recall list.

Sixty percentage (n=346) of recalled pharmaceutical products were modern or allopathic (208) and 35% were traditional or ayurvedic (120) (figure 1B). Two-sided fisher exact test showed that significantly large number of modern pharmaceutical products were recalled (p-value< 0.001). Twenty-seven percentage of the recalled drugs were unregistered at the DDA indicating they were not authorized to distribute and sell in Nepal. Similarly, twenty percentage of the recalled drugs, mostly allopathic, were listed as essential medicines and have been distributed free of cost through government health centers. Remaining 40% were non-essential ones (p-value < 0.01) and equal number of drugs were categorized as others, mostly ayurvedic since such drugs are not classified as essential or non-essential. Most of the recalled pharmaceutical products were substandard (215) followed by unregistered (93) and falsified (38) (see figure 1B). We found that the recall pattern among these three categories were significantly different (one-way chi-square test, p-value < 0.001, X-squared = 118.66, df = 2). The recalled pharmaceutical products included a significantly (two-sided fisher exact test, p-value <0.05) higher number of domestically manufactured (134) items than the imported (107) ones which were manufactured mostly in India (figure 1B). Few drugs from Bangladesh and China were also recalled. Country of origin of 105 recalled pharmaceutical products were not identified.

Based on the generic names of each non-ayurvedic pharmaceutical product, we identified their generic names and then categorized them into different groups based on their therapeutic properties. The top 10 most recalled drugs belonged to antimicrobials (46) followed by gastrointestinal medicines (35), vitamins and minerals (29), antiseptic (23), hormones and contraceptives (18), and pain and palliative care medicines (16), fluid and electrolyte replenishment (13), cardiovascular and renal drugs (7), anti-diabetes (5) and antihistamines (5) (see figure 2A). Remaining recalled drugs were CNS drugs, respiratory system drugs, prostaglandin analogues, antirheumatic. Nineteen drugs were not classified into any of those and labeled as “others” since not enough information was available. The DDA provided reason(s) for every recalled pharmaceutical product. Most of the drugs were recalled because they were not registered (93) at DDA. The most common reason for recall among registered drugs was the failure to comply with microbial test (82) followed by failures in dissolution test (40), in quantitative assay for active pharmaceutical ingredient (21), and in physical characteristics (17). Eleven products did not comply label requirements and 12 had one or more impurity. others (see figure 2B). Tablets were the most recalled dosages form followed by powder, solution, capsules, syrups/suspension, cream/ointment and other (figure 2C). The shelf-life of recalled drugs ranged from less than three months (16.4%) to more than two years at the time of recall (figure 2D).

**Figure 2:**
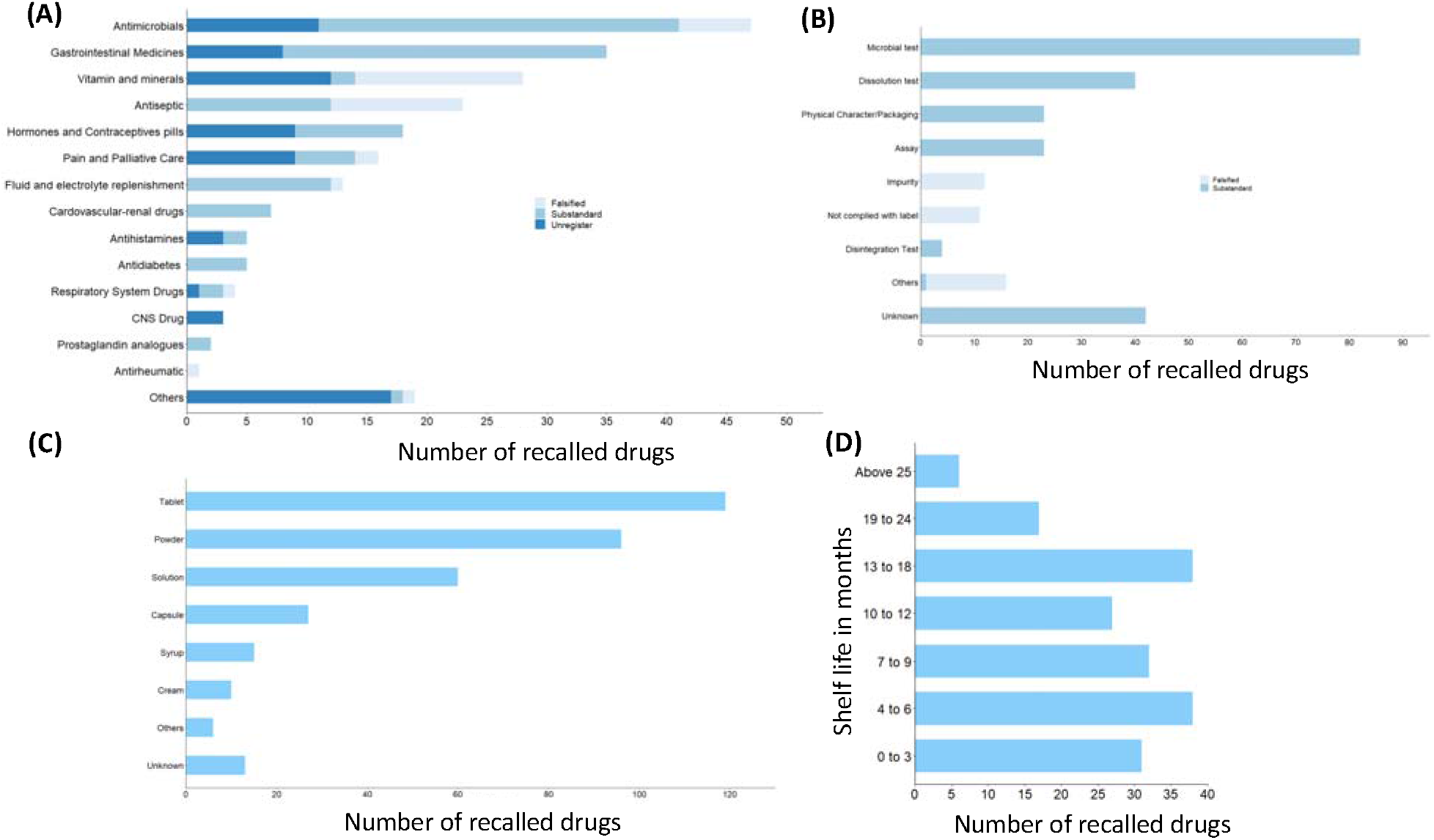
(A) Different categories of recalled drugs based on their therapeutics, (B) List of major reasons for recalling pharmaceutical products, (C) Types of dosage forms of recalled drugs, (D) Self life of recalled pharmaceutical products after the recall (in months).

### Low-quality drugs reported in research papers

We also systematically looked into published research works in order to find the reporting of low-quality drugs in Nepali market. Initially, we identified 467 journal articles through the literature search in four different databases: PubMed, Springer link, Web of science and Google scholar. We removed 13 duplicate articles and brought the number of articles to 454. By screening the title and abstract of these articles, we removed 439 articles and we considered only 15 in next step. We read the full text of these articles and excluded 11 articles because they did not follow the inclusion criteria. At last, four articles were found to be relevant that contained primary information on the prevalence of substandard, falsified, and unregistered medicines in the Nepali market (list is given in supplementary information). The flow chart of the study is given in a flow chart in figure 3.

**Figure 3:**
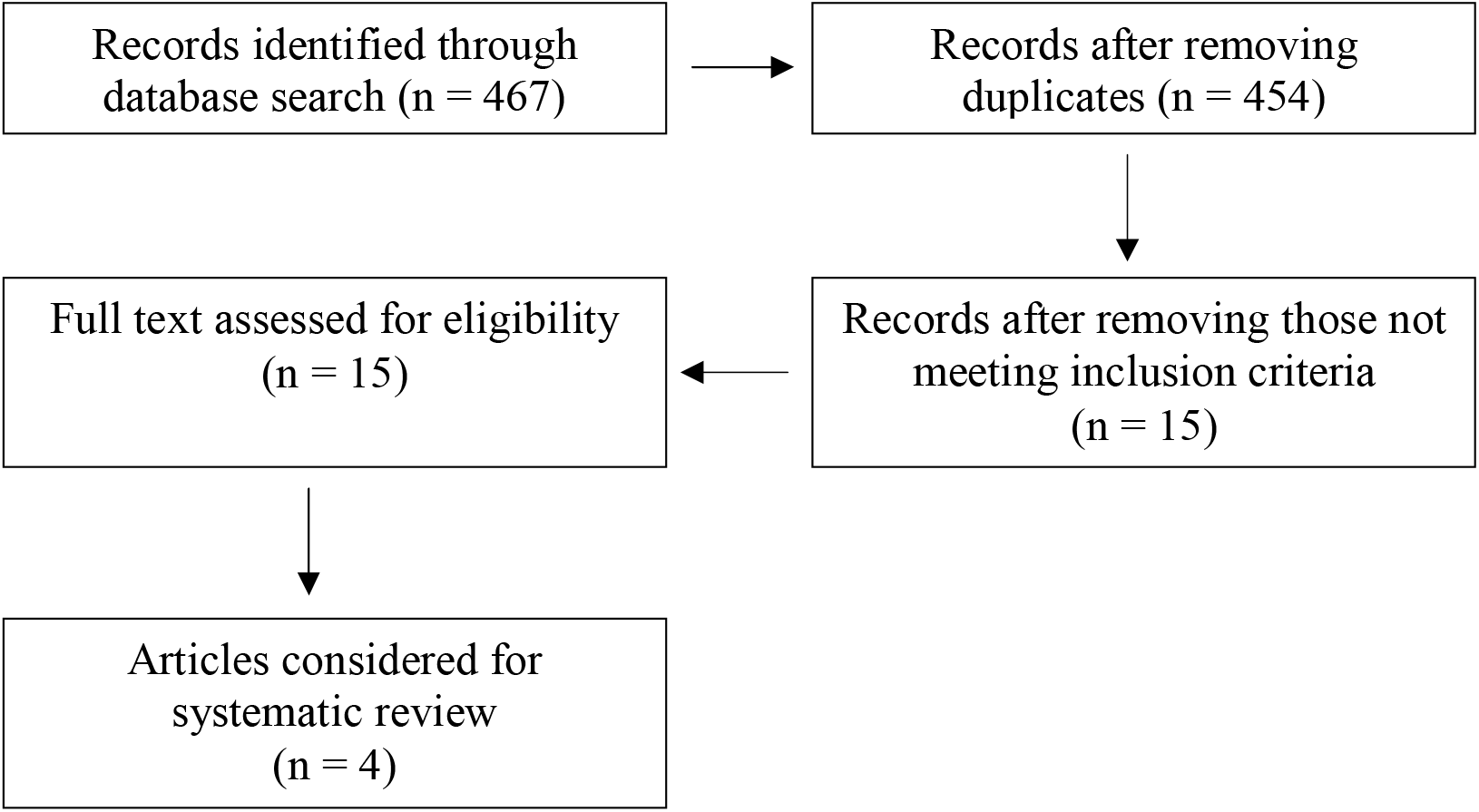
Flow chart of research papers search procedure

A cross sectional descriptive study reported by Jha et al.^13^ assessed the quality of essential medicines available in public health care facilities of Nepal. The study was carried out in 62 health facilities in 21 districts of Nepal representing all seven provinces. Out of 244 batches of 20 different generics of essential medicines tested, 37 batches failed to meet the required pharmacopeial standards which constitute 62.16% batches of medicines supplied by Government of Nepal and 37.83% batches purchased from local resources. The failed medicines included antibiotics, supplements, anti-diabetics etc. Providing required information on the label is another issue. Majority of the pharmaceutical products inspected in Chitwan in 2017 involving 759 drugs from 37 Nepali pharmaceutical companies did not meet the regulatory standards of primary labeling^8^. Most of them missed at least one critical information on the label such as drug quantity, name of pharmacopoeia, serial number of pharmaceutical industries, price list, drug classification, and information in Nepali language. Majority of the products (84%) did not provide the directions of use. Similarly 90% of drug samples (n=40) in Kathmandu did not comply with the existing regulatory requirement on labeling and 42.5% brands did not mention about the pharmacopoeial standard^7^. The same study showed that 40% of domestic and 28% imported brands failed to meet national criteria during laboratory analysis. In average 32.5% samples were found to be of substandard quality in this study. This study aims to evaluate the availability and rationality of unregistered fixed-dose drug combinations (FDCs) in Nepal. A snowball sampling method with visits to 20 retail pharmacies in each of five major cities in Nepal was used to assess the availability of unregistered FDCs. To justify the rationality of the FDCs obtained from these five cities, the toolkit developed by Health Action International Asia-Pacific (HAI-AP) was used. Forty-one unregistered fixed-dose drug combinations were found in five major cities of Nepal, majority of which were anti-inflammatory/analgesic/antipyretics. Regulatory authorities should initiate strict monitoring and appropriate regulatory mechanisms to prohibit the use of unregistered and irrational FDCs.^9^

## Discussion

The low quality medicines are recalled from the market by manufacturing company voluntarily or by the order of national or international drug regulatory bodies to protect the public from a defective or potentially harmful product^14^. Many recall incidents of poor quality medicine have been reported globally^15^. In 2010, Johnson and Johnson recalled the drug which was deviated from good manufacturing practices (GMPs). Similarly, in 2012, the US FDA recalled the contaminated vials of corticosteroid medication which was manufactured by the New England Compounding Center^16^.

The overall trend of recalled drugs is increasing in Nepal. Starting from a single drug recall in 2010 to highest numbers (123) in 2018. In this year, most of the drugs (90) were recalled since they were not registered with the DDA. This indicates that the circulation of unregistered drugs is a serious issue in Nepal which may been contributed by the open and unregulated boarder with India. Both allopathic and ayurvedic medicines are widely used in Nepal. The allopathic medicines are the modern medicines that are manufactured synthetically whereas ayurvedic medicines are the traditional medicines which uses the natural remedies to improve health or to treat diseases. Both types of medicines are commercially manufactured in Nepal in addition to be imported mostly from India. There are two groups of manufacturers of ayurvedic drugs in Nepal. Mostly they are manufactured by registered commercial companies and sold in market in packages through registered shops. The ayurvedic drugs are also manufactured by individuals or small business holders who may not be registered at DDA. They sell their ayurvedic products in streets, through door-to-door service, and through individual networks. We found that both allopathic and ayurveda medicines were recalled due to their non-compliance with government standards. Ayurvedic medicines are utilized prominently in Nepalese communities, and sometimes, there are used concomitantly with allopathic^17^. There has been an increasing interest in the study of traditional medicine in different parts of world^18^. However, there is the lack of quality research and standards, and stringent regulatory environment in this sector. Hence, the successful implementation of herbal pharmacovigilance, GMP as well as inspection on herbal medicine formulation is essential for developing reliable information on the safety of ayurveda medicines^17^.

Essential medicines are defined by WHO as the medicines that satisfy the priority healthcare needs of the population. In Nepal, the concept of Essential medicines was adopted in 1986 A.D to enhance the access of essential medicines to every individual. The main criteria for selection of the medicines in the National List of Essential Medicine (NLEM) are public health relevance, efficacy, safety, cost-effectiveness and access of the drugs. The NLEM 2016 of Nepal contains 359 medicines which has 86 medicines more than the NLEM 2011^10^. Following criteria were used for including a medicine in NLEM: approved and licensed in Nepal, relevance to a disease posing public health problem, proven efficacy and safety, aligned with standard treatment guideline of Nepal, stable under storage conditions, cost-effectiveness, access. However in following conditions medicines were excluded from the NLEM list: banned in Nepal, safety concerns, if medicine with higher efficiency, safety profile and lower-cost is available, irrelevant to public health disease burden, antimicrobial resistant, medicine with abuse and misuse potential^10^. Our study shows that some of the recalled allopathic medicines also listed under National essential drug list by the Government of Nepal. Jha et.al^19^ indicated the presence of high number of substandard essential medicine and majority of which were purchased by Government of Nepal. Essential medicines for various illnesses are supplied free of cost in Nepal through government hospitals, health care centers and health posts. Poor quality of essential medicines can have serious impact on public health. As significant proportion of drugs recalled by DDA included essential medicines distributed by Government of Nepal, there is enough room to improve the procurement procedure and upgrading of health facilities of Nepal that store and distribute the medicines. In one study^20^ that looked into the procurement practices in Nepal reported that the majority of hospital pharmacies in Nepal use an expensive direct-procurement model for purchasing medicines. They relied on on doctors’ prescriptions to choose a particular brand, which may be influenced by pharmaceutical companies’ marketing strategies. Most of the hospital pharmacies procured only registered medicines, a minority reported purchasing unregistered medicines through unauthorised supply-chains. Not all pharmacies followed Basel Statements during procurement of medicines. Such pharmacies may need awareness and training to fully adopt regulation of national and international policies for enhancing accessibility to quality medicines.

Among the recalled group, antimicrobial group of medicines had the highest frequency of incidents. Acharya et.al^21^ highlighted the problem of antimicrobial resistance in Nepal as an alarm bell for an even worse public health situation. Suboptimal dose or poorly manufactured antibiotic medicine increases the chance of antimicrobial resistance which is an emerging health menace in current situation^22^. Most of the recalled therapeutic categories of medicines like vitamins and minerals, NSAID, antipyretic and analgesic, antiseptic, fluid and electrolyte replenishment and others are over-the-counter medicine that can be brought from the pharmacy without the prescriptions and such medicines can pose a significant threat to the groups of patients who consume them^23^. Similarly, few antidiabetes medicines were also recalled. Consumption of such medicines will increase the incidence of macrovascular and microvascular complications due to compromised glucose control^24^.

Our study shows that the drugs were recalled due failure in various laboratory tests like assay, content uniformity test, weight variation, impurity test, dissolution test, frability test, identification and sterility test. This phenomenon might be attributed to the lack of proper quality control during manufacturing and lack of following proper procedures for transportation and storage conditions^12^. Jha et.al pointed out that recommended range for medicine storage in some selected health facilities in Nepal was highly ignored^19^. Keeping the temperature and humidity within a range is must necessary because it has a major role in degradation of medicines. Another reason was failure to comply with the claim and incorrect labeling. The DDA regulation requires appropriate labeling of marketed medicines to ensure patient medication safety, which seems to be not followed properly. Thus, the drug analyst and the drug regulators should be encouraged to remain vigilant about counterfeiting possibility and conduct the analysis including chemical, physical, package inspection, and authentication efforts to determine quality more accurately^25^.

Domestically produced and imported medicines in Nepal should have the registration license from DDA^6^. Nonetheless, we found that numbers of unregistered drugs were recalled during the inspection. The drug supplier, whole seller, and retailers should ensure that the drug is registered within the national regulatory body to timely identify substandard products before they reach patients. Also, the regulatory body should stringent post-market surveillance to ameliorate the situation. The unregistered medical products in Nepal may or may not have been registered in India. Since Nepal shares open and poorly regulated boarder with India, drugs registered in India are also easily sold in Nepali market. We found that nearly half of the total recalled medicines were imported from India. India is the leading country in counterfeit drug production, having as much as 35% of the world production originating within its borders^26^.

One of the main reasons for the circulation of low-quality drugs in low-income countries is the improper storage and transportation conditions. One study indicated only 13% of health facilities (n=62) were found to follow the medicine storage guidelines in Nepal regarding sunlight protection, humidity protection, heat protection and maintenance of ventilation^13^.

## COVID-19 related recalls

The COVID-19 pandemic has resulted in the surge of substandard and falsified medical products including drugs, masks, sanitizers, diagnostic tests and vaccines not just only related to COVID-19 but other essential medical products^27^. Rampant circulation of fake medical products during emergencies has happened throughout the history^27^. Counterfeit respirators and masks pose additional risk to health care workers^28^. Falsified chloroquine was seized in Cameroon, Congo and Niger during March and May 2020. Chloroquine was controversially announced as the drug for the treatment of COVID-19^29^. Similarly, in 2019, with an outbreak of Covid-19 pandemic, the FDA has uncovered nearly 1,300 fraudulent products^30^. With the outbreak of Covid-19, in March 2020, DDA Nepal has amended the standard for Instant Hand Sanitizer in order to prohibit the selling of the substandard, falsified and unregistered sanitizers^31^. In between September and November 2020, the DDA issued the recalled notice for 19 hand sanitizers which failed to comply with the standard guideline. Some sanitizers were found to contain methanol, rather than ethyl alcohol and isopropyl alcohol. As methanol is very toxic, some of the case series indicated use of hand sanitizer containing methanol causes the transdermal absorption and increases the risk of systemic toxicity^32^. The increase in the demand of hand sanitizer and other medicines has increased the growth of e-commerce. Online sale of pharmaceutical products has just started in Nepal during recent years. WHO has reported that 60% of medications purchased through Internet could be counterfeit or substandard, and more than 50% of medications purchased online from sites that concealed their actual physical address was found to be low quality medicine^33^. Nepali regulating agencies should pay special attention to this new method of business in Nepal to protect people from the consumption of low-quality and fake medical products. Inexorable growth of online pharmacies, unregulated websites, and, social media platform for business may contribute to the dispensing of unapproved, subpotent, counterfeit, expired or illegal drugs, and prescription drugs without a valid prescription in Nepal too^34^.

## Conclusion

In this paper we presented a detailed analysis of low-quality and fake drugs circulating in Nepal in the past decade using recall notice of DDA Nepal. We showed that the number of recalled drugs has significantly increased. This might be attributed to greater surveillance by DDA or the substandard, falsified and unregistered medicine in the market are actually increasing. Similar to global trend, antimicrobial drugs were the most recalled drugs in Nepal. Since antibiotics are available over the counter in Nepal without doctor’s prescription, it is necessary to enforce strict regulation so that the rampant (mis)use of such drugs is minimized and prevent antibiotic resistance. We relied on recall notice from DDA. The recall notice does not provide information on the number of samples collected for testing and location of sample collection. Therefore, our analysis did not report the rate or prevalence of low-quality drugs. Since sample collection locations were not available, it was not possible to know the most vulnerable regions of Nepal for low-quality drugs. Therefore, a systematic study is needed to understand the prevalence of substandard and falsified drugs in Nepal covering different parts of the country on regular basis. We suggest having more stringent regulatory systems and implementation for pharmaceutical manufacturing industries and post marketing surveillance.

## Data Availability

All data included in the manuscript and supplementary information.

