## Supplementary material for "Incidences of poor-quality pharmaceutical products in Nepal": SI Table 1

Table SI1: Result of literature search

| Keywords | Number of articles found | | |  |
| --- | --- | --- | --- | --- |
|  | PubMed | Web of Science | Springer link | Google Scholar |
| Counterfeit* OR substandard* OR fake OR spurious OR unregulated OR unregistered OR falsified* OR fraud | 14532 | 51113 | 71770 | 41100 |
| Drug OR medicine OR pharmaceutical | 4977168 | 4579007 | 1386455 | 728000 |
| Nepal* | 12868 | 18132 | 16862 | 26500 |
| 1 AND 2 AND 3 | 13 | 10 | 393 | 51 |

**List of research papers that included drug quality in Nepal**

1. Poudel, Ramesh Sharma, et al. "Assessment of primary labeling of medicines manufactured by Nepalese pharmaceutical industries." *Journal of pharmaceutical policy and practice* 11.1 (2018): 1-6.
2. Gyanwali, P., et al. "Surveillance of Quality of Medicines Available in the Nepalese Market: A Study from Kathmandu Valley." *Journal of Nepal Health Research Council* (2015).
3. Poudel, Arjun, et al. "Assessment of the availability and rationality of unregistered fixed dose drug combinations in Nepal: a multicenter cross-sectional study." *Global health research and policy* 2.1 (2017): 1-13.
4. Jha, A. K., et al. *Quality of essential medicines in public health care facilities of Nepal–2019*. Nepal Health Research Council, 2019.
